# Fibrinogen-Associated Plasma Metabolites and Implications for Coagulation, Inflammation, and Vascular Diseases

**DOI:** 10.1101/2025.08.06.25333064

**Authors:** Jayna C. Nicholas, Taryn Alkis, Joshua C. Bis, Eric Boerwinkle, Jennifer A. Brody, Clary B. Clish, Paul S. de Vries, Yan Gao, Robert E. Gerzsten, Xiuqing Guo, Andrew D. Johnson, Martin G. Larson, Rozenn N. Lemaitre, Bruce M. Psaty, Vasan Ramachandran, Alexander P. Reiner, Stephen S. Rich, Benjamin Rodriguez, Jian Rong, Jerome I. Rotter, Jeanette Simino, Nicholas L. Smith, James Wilson, Jie Yao, Alanna C. Morrison, Bing Yu, Laura M. Raffield

## Abstract

**Background:** Fibrinogen is a critical coagulation factor that plays an essential role in thrombosis and is elevated in individuals with chronic inflammation. Here, we used fibrinogen as a representative quantitative measure of pro-coagulant risk and evaluated metabolites associated with fibrinogen levels through non-targeted plasma metabolomic profiling (Broad and Metabolon platforms).

**Methods:** Our analysis included 10,533 individuals across six U.S. based cohorts representing diverse population groups. The cross-sectional relationship between each of 789 tested metabolites and plasma fibrinogen concentration was assessed with adjustment for relevant covariates such as age, sex, body mass index, and circulating lipoprotein levels.

**Results:** Meta-analysis of per-cohort results revealed 270 metabolites significantly associated with fibrinogen level (FDR adjusted p-value < 0.05). Lipid species such as glycerophospholipids, sphingolipids, and fatty acyls were prevalent among significantly associated metabolites; some of these may capture effects of inflammation, as supported by sensitivity analyses adjusted for C-reactive protein. Significant associations between fibrinogen levels and serotonin, thyroxine, and sex-hormone derivatives may capture endogenous influences on fibrinogen levels. Exogenous compounds and microbial co-metabolites significantly associated with fibrinogen also implicate lifestyle and microbiome risk-factors. Only a portion of fibrinogen-associated metabolites (30%) have been associated with a cardiovascular disease outcome in a prior study, suggesting the associations discovered here may provide insights on vascular biology which case-control studies may not yet be powered to detect.

**Conclusions:** These findings contribute to a growing list of metabolite biomarkers that may influence coagulation and inflammation pathways and may thereby contribute to vascular risk.

## Introduction

The coagulation cascade and acute-phase inflammatory response are intertwined in pathogenesis of adverse vascular conditions such as atherosclerosis and thrombosis. Fibrinogen is a circulating coagulation factor and acute phase reactive protein which is constitutively expressed and upregulated in response to interleukin-1 (IL-1) and interleukin-6 (IL-6) mediated inflammatory pathways (1). Although it is unclear whether fibrinogen has a causal effect on vascular risk in human populations(2,3), higher levels robustly associate with cardiovascular disease (CVD) -related phenotypes including increased blood viscosity, thrombosis, atherosclerosis(4), and incident coronary heart disease (CHD), ischemic stroke, and venous thromboembolism (VTE) risk(5,6).

Regulation of circulating fibrinogen levels is multi-faceted, involving both heritable and environmental contributions. Baseline levels of circulating fibrinogen vary across individuals and populations, with higher levels generally observed among African Americans (AA)(7) and female(8) participants. While genetic variants (9)(10) (11) (12) (13) and some modifiable lifestyle factors such as physical activity and smoking(14) are known to associate with circulating fibrinogen levels, the pathways regulating fibrinogen are not fully understood and likely have uncharacterized mediators.

Metabolites are active players in the coagulation and inflammation pathways – with, for example, vitamin K as a critical co-factor for post-translational modification of coagulation and anticoagulation proteins required for their functional activity (15), and prostacyclin serving as a lipid inhibitor of platelet aggregation(16). Furthermore, metabolites may mediate and/or mark disease progression, as seen with cholesterol’s role in atherosclerosis and cardiovascular disease(17). As an individual’s ‘metabolome’ captures both endogenous and exogenous influences, studies of metabolites may reveal insight on internal and external environmental molecular contributors to disease-relevant processes and highlight new interventions. In recent years, advances in metabolomics have provided the opportunity to study the relationship between thousands of metabolites and disease or disease-related pathways. To date, most metabolomics studies have focused on metabolite associations with disease endpoints. However, associations between metabolite levels and quantitative measures reflecting relevant pathways may provide new insights on the molecular basis of disease(18–21). For example, a 2017 study associating plasma metabolites with circulating fibrinogen measures in 925 European participants highlighted 36 metabolite associations thought to represent inflammatory and lifestyle or dietary influences (19).

Here, we assessed plasma/serum metabolite associations with fibrinogen using untargeted metabolomics data from six U.S.-based cohorts (total n=10,533) representing diverse population groups. Given fibrinogen’s position at the crossroads of coagulation and acute-phase inflammation pathways (**Figure 1**) (22), we hypothesize that metabolites levels significantly associated with fibrinogen measures may capture markers or drivers of thrombotic and inflammatory risk, and potentially influence downstream vascular phenotypes.

**Figure 1.**
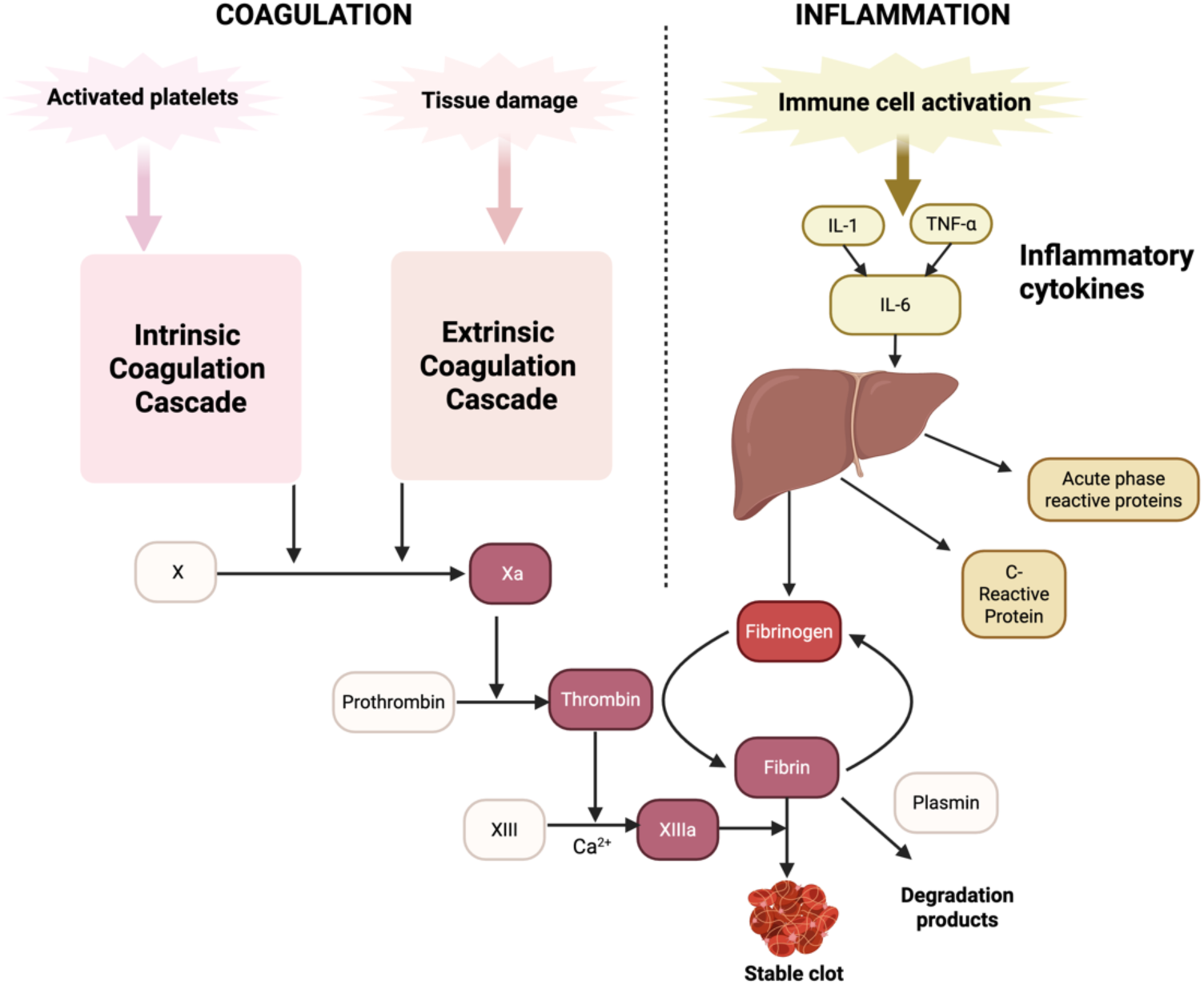
Graphical depiction of coagulation and inflammation pathways. Fibrinogen stands at the cross-section of these pathways and is influenced by both.

## Results

### Basic Characteristics of Study Population

In total, 10,533 participants were included in the present study. Our study included 5,509 participants self-identifying as Non-Hispanic White (NHW) (ARIC: 1,475; CHS: 322; FHSII: 2,316; FHSIII: 975; MESA: 421); 4,719 self-identifying as Black or African American (ARIC: 2,273; JHS: 2,251; MESA: 195); and 305 self-identifying as Hispanic American (MESA: 305) (**Table 1**). Details of participants from each individual study are provided in **Supplemental Table 1**.

### Overview of significant metabolite associations with fibrinogen

In total, 789 metabolites were measured in at least two cohorts with missingness less than or equal-to 25% and were thus tested in meta-analysis. Of these, 270 metabolites were significantly associated with plasma/serum fibrinogen level, as measured by Clauss assay or immunonephelometric assays (FDR-adjusted p-value<0.05) (**Table 2; Supplemental Table 2; Figure 2**). Of the significantly associated metabolites, 152 showed positive associations and 118 showed inverse associations indicating a roughly equal distribution of effect directions. All 9 plasma metabolites that were significantly associated with fibrinogen in prior studies (19,20) and tested in the present meta-analysis were significantly associated with fibrinogen with concordant directions of effect here (**Supplemental Table 5**). A total of 261 metabolite associations observed here were not, to our knowledge, observed in a prior study.

**Figure 2.**
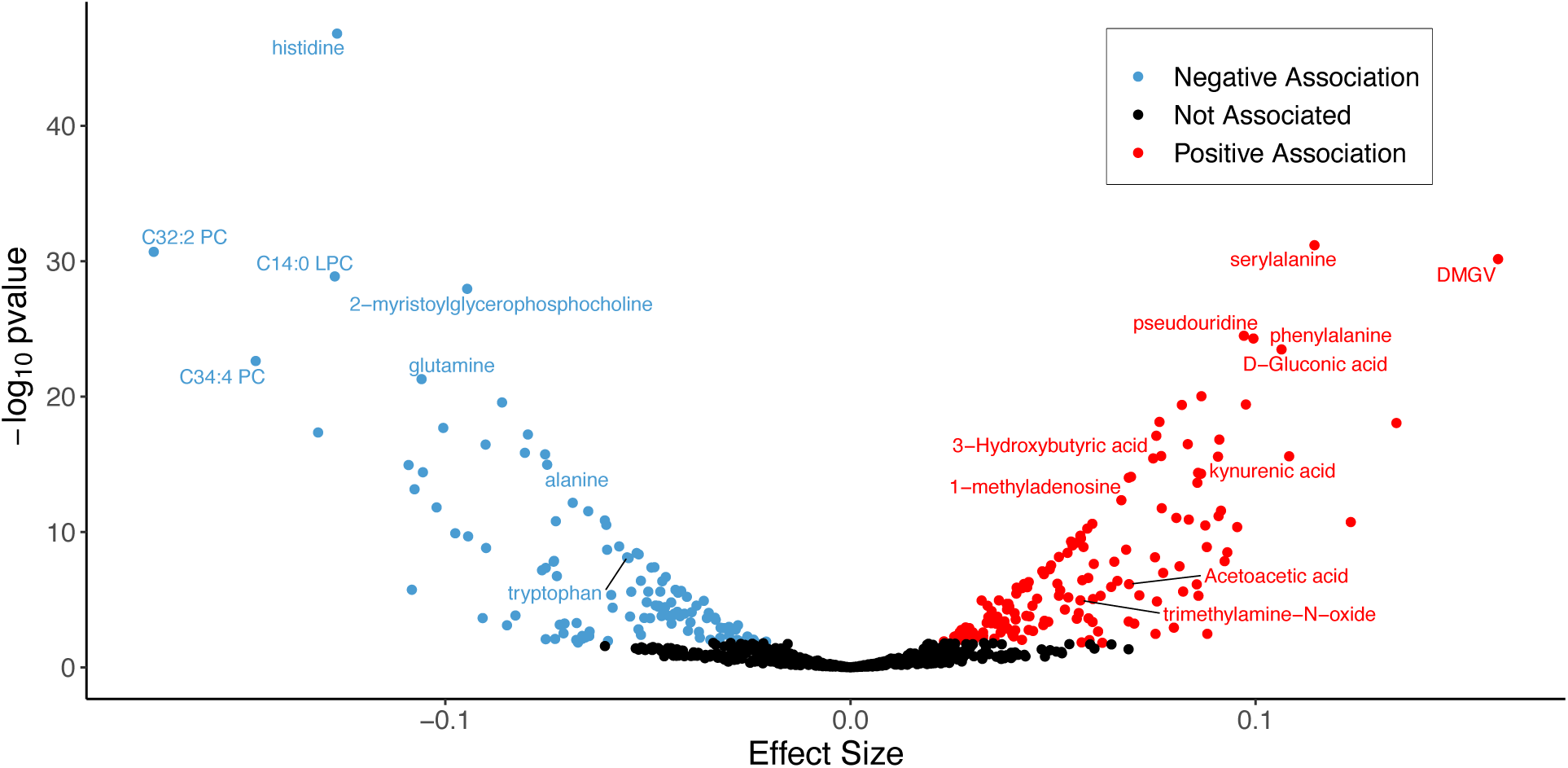
Volcano plot of effect sizes obtained from meta-analysis assessing metabolite associations with circulating fibrinogen. 152 positive effect-size and 118 negative effect-size associations were observed.

Lipids, organic acids, and organic heterocyclic compounds were the most prevalent super-classes represented among fibrinogen-associated metabolites - accounting for 48% (132), 31% (84), and 8.14% (22) of significant metabolites, respectively (**Figure 3A**). Metabolites classified in the HMDB “organic acids and derivatives” super-class were statistically overrepresented (Fisher’s Exact Test FDR-adjusted p=0.026) among significant metabolites compared to metabolites tested in the meta-analysis. A positive association between fibrinogen and peptides with clear biological links to fibrinogen or coagulation, such as Fibrinopeptide A degradation product DSGEGDFXAEGGGVR, and Complement Protein C3b degradation products (HWESASLLR, HWESASXX), reflect known correlation between the complement and the coagulation pathways and suggest that we are capturing on-target associations. An inverse association between histidine and fibrinogen was the most significant result meta-analysis wide (β = - 0.18, SE = 0.01, FDR-adjusted p-value = 1.6 𝑥 10^-47^), while dipeptide serylalanine was the most significant positive association (β = 0.12, SE = 0.01, FDR-adjusted p-value = 1.96 𝑥 10^-29^). Sex-stratified analyses revealed 11 metabolites significantly associated with fibrinogen levels in only males (not in meta-analysis or females-only), and 18 metabolites significantly associated with fibrinogen levels in females (not in meta-analysis or males-only) (**Supplemental Table 11 (Male Secondary Results); Supplemental Table 12 (Female Secondary Results), Supplemental Figure 2**). Significant effect-size heterogeneity (i.e. Cochran’s Q p-value<0.05) between sexes was observed for 17 of these 35 metabolites.

**Figure 3.**
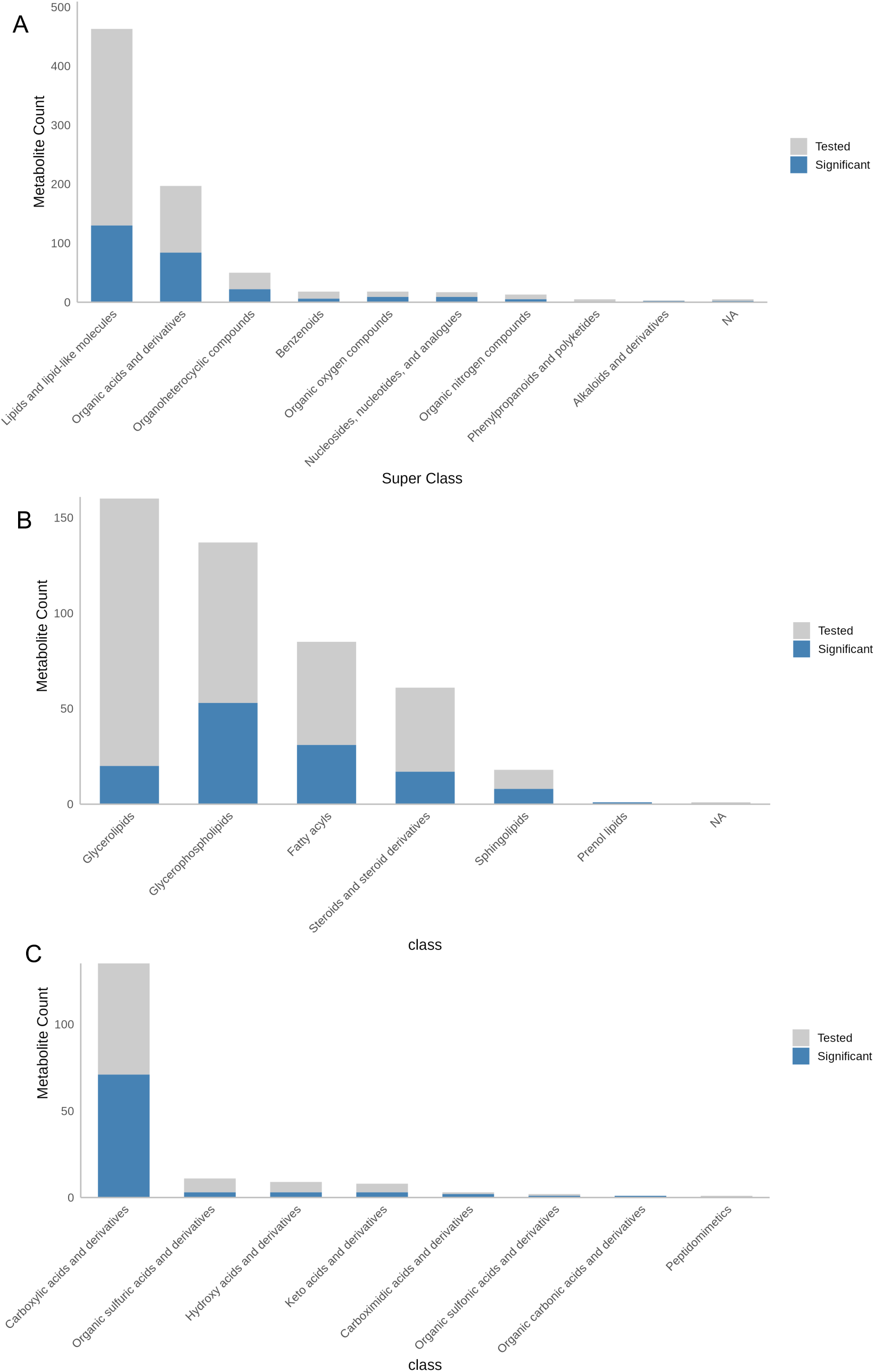
(A) Proportion of tested metabolites in each Human Metabolome Database (HMDB) superclass significant in meta-analysis. Lipids comprise the largest percentage of meta-analysis significant metabolites. Organic acids and derivatives are statistically enriched among significant metabolites compared to tested. (B) Lipids and (C) organic acids in each HMDB class tested (gray) and significant (blue) in the meta-analysis. Y-axis scales differ by plot.

**Figure 4.**
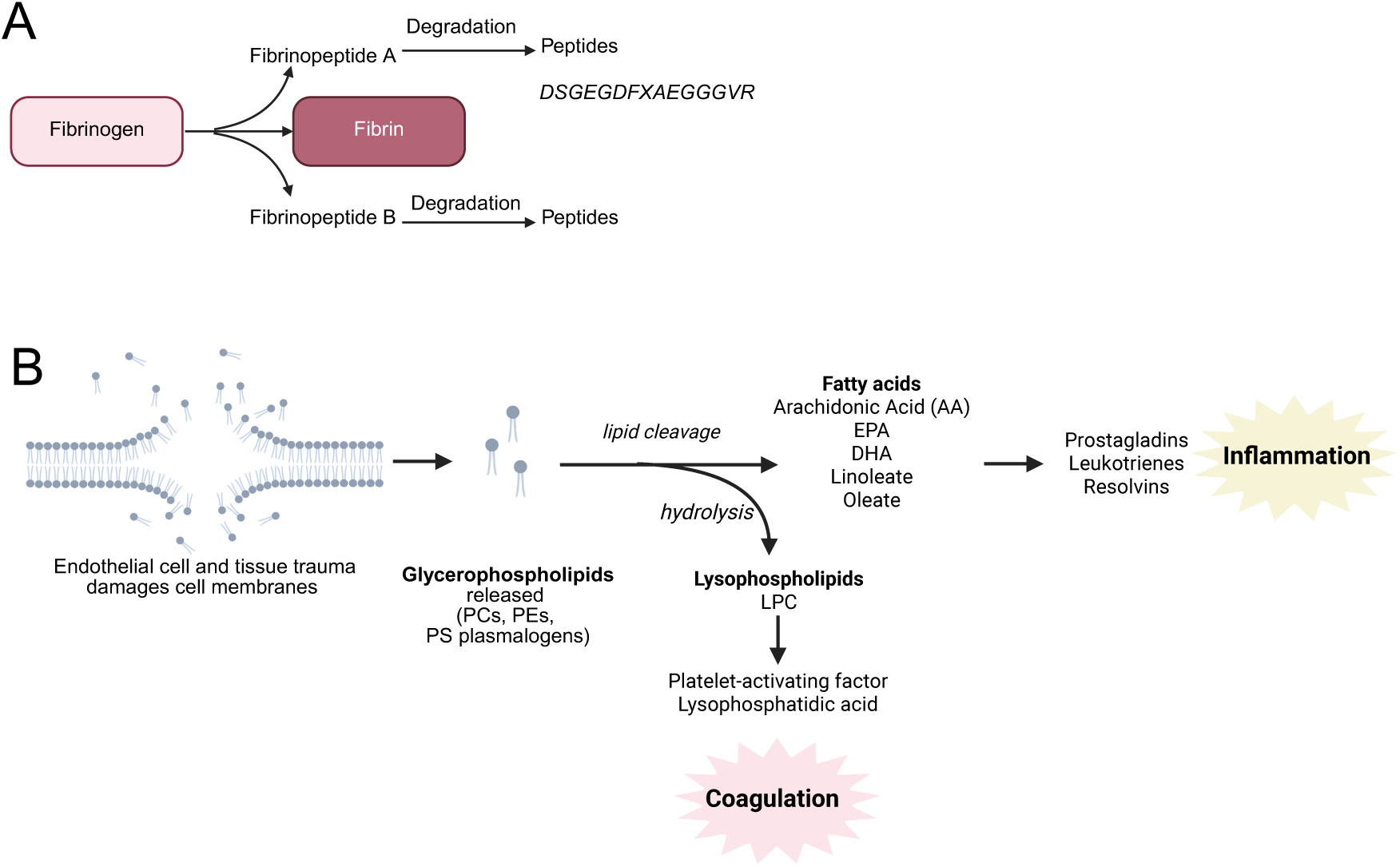
(A) Graphical depiction of fibrinopeptide A generation. Fibrinogen cleavage yields fibrinopeptides A and B. Fibrinopeptide A is cleaved to form the peptide derivative *DSGEGDFXAEGGGVR,* which is positively associated with fibrinogen in the present study. (B) Graphical depiction relating glycerophospholipids, fatty acids, and lysophospholipids, which were significantly associated with fibrinogen in the meta-analysis, to pathways of coagulation and inflammation based on known relationships in literature.

### Fibrinogen-associated metabolites capture inflammation-related changes in metabolome

The majority of fibrinogen-associated metabolites were lipids (**Figure 3A, 3C**) which may capture inflammation-relevant metabolite alterations. Glycerophospholipids were the most prevalent class of lipids significantly associated with fibrinogen (**Figure 2B**) and were enriched for inverse associations (47/53, binomial test p-value p=5.8 𝑥 10^-9^, FDR-adjusted p-value=1.74 𝑥 10^-7^, **Supplemental Table 15, Supplemental Figure 3**). Omega-3 polyunsaturated fatty acids arachidonic acid (AA), eicosapentaenoic acid (EPA), and docosahexaenoate (DHA) were inversely associated with fibrinogen (**Supplemental Table 15, Supplemental Figure 3)**. Fatty acyls were nominally enriched for positive associations with fibrinogen (23/31, X^2^ binomial test p-value=1.1 𝑥 10^-2^, FDR-adjusted p-value =0.11, **Supplemental Table 15, Supplemental Figure 4**); several of these metabolites (16/23, 70%) were acyl carnitines.

Sphingolipids were also nominally enriched for positive associations (8/8, X^2^ binomial test p-value=7.8x10^-3^, FDR-adjusted p-value=0.11, **Supplemental Table 15, Supplemental Figure 5**). Positive associations between fibrinogen and amino acids (**Supplemental Figure 6)**, nucleotides/nucleosides, as well as inverse associations with heme degradation products biliverdin, bilirubin (Z, Z), and bilirubin (E,E) may also capture metabolomic signatures of inflammation.

C-Reactive Protein (CRP) is another acute phase reactive protein secreted from the liver in response to IL-6 stimulation and is a marker of inflammation. Of the fibrinogen-associated metabolites identified, 33 have been associated with CRP in a prior study (30/33 with concordant directions of effect - i.e. higher CRP with higher fibrinogen)(19,20) (**Supplemental Table 6**). Additionally, 9 metabolites have previously been associated with white blood cell count (WBC), and 11 previously associated with interleukin-6 (IL-6), which are both considered indicators of inflammation as well (19,20)(21). To further assess which of the fibrinogen-associated metabolites may be influenced primarily through inflammatory pathways, we performed a sensitivity analysis adjusting for CRP in JHS, which is one of the large cohorts with CRP and fibrinogen measured at the same time point (this was not available across all cohorts). Of the 129 metabolites significantly associated with fibrinogen in the meta-analysis (FDR p-value<0.05) and nominally-associated in JHS (JHS p-value <0.05) prior to CRP adjustment, 6 metabolites were attenuated to non-significance (JHS p-value < 0.05, CRP-adjusted JHS p-value > 0.05, p-value fold-increase >=2, effect size decrease >=50%) following adjustment for CRP (**Table 3, Supplemental Table 9, Supplemental Figure 7**). Three of these attenuated metabolites were lipids - C18:1 lysophosphatidylcholine (LPC), C18:1 lysophosphatidylethanolamine (LPE), and C20:4 LPC - while the other three were organic acids - N-acetylputrescine, serine, and threonine. Eleven additional meta-analysis significant metabolites were attenuated towards non-significance by CRP-adjustment in JHS, including LPC, lysophosphoethanolamine (LPE), sphingomyelin species, as well as amino acids arginine, asparagine, proline, and GABA (**Table 3**). Finally, a subset of 11 additional metabolites - including lipid species C20:0 LPE, C14:0 carnitine, and amino acids glycine, citrulline, N-acetylhistidine - were only significant in JHS after adjustment for CRP (JHS p-value>0.05, CRP-adjusted JHS p-value < 0.05, p- value fold-increase >=2, effect size increase >=50%) (**Table 3**). However, most fibrinogen associations were robust to CRP adjustment, suggesting there may be distinct metabolic signatures of fibrinogen versus CRP.

### Metabolite associations reveal putative endogenous influences on fibrinogen

We note several hormone metabolites significantly associated with fibrinogen which may capture endogenous pathways influencing, or altered in parallel to, fibrinogen levels. Steroid-fibrinogen associations included cortisol, as well as 8 androgen and progesterone derivatives. Although these metabolites are involved in sex hormone biosynthesis and degradation, none of these associations were exclusive to males or females or showed strong evidence of effect size heterogeneity by sex (I^2^<25% for these metabolites, **Supplemental Table 2**). Three amino-acid derived hormones were positively associated with fibrinogen - including tryptophan-derived serotonin, gamma aminobutyric acid (GABA), and tyrosine-derived thyroxine. GABA biosynthetic pathway components N-acetyl putrescine, 4- acetamidobutanoate, and 4-guanidinobutanoate were also positively associated with fibrinogen.

Kidney function can be impaired in individuals with high cardiometabolic disease risk and is a strong driver of endogenous metabolite levels. In literature, fibrinogen levels show inverse correlation with estimated glomerular filtration rate (eGFR), and may be an independent risk factor for chronic kidney disease (23). To assess which of the present fibrinogen-associated metabolites may be influenced primarily by kidney function, we performed a sensitivity analysis adjusting for eGFR in ARIC-African American (n=1,929), as well as ARIC NHW (ARIC-NHW, n=1,475) cohorts. Of the 139 metabolites nominally associated with fibrinogen in both ARIC cohorts (p<0.05) and significantly associated in the meta-analysis, 2 metabolite associations (2-methylmalonyl carnitine, deoxycarnitine) became non-significant in both cohorts following adjustment for eGFR (ARIC p-value <0.05, eGFR-adjusted p- value>0.05, p-value fold increase >2) in ARIC post-eGFR adjustment; however, the effect sizes remained within 50% of original value (**Supplemental Table 10**). This suggests that metabolite- fibrinogen associations observed may be relatively robust to influences of eGFR.

### Exogenous compounds reveal possible environmental influences on fibrinogen

Several environmental exposures, such as smoking and dietary consumption of red meat, are known to correlate with levels of circulating fibrinogen and increase risk of cardiovascular disease outcomes (24)(25). Accordingly, we found tobacco metabolite cotinine, as well as caffeine and vitamin A, were among significant metabolites positively associated with fibrinogen. Diet-derived compounds, choline and betaine (which are often derived from animal products), and downstream gut microbiota metabolite trimethylamine N-oxide (TMAO) were also significantly, positively correlated with fibrinogen levels. Metabolites previously linked to coffee consumption - including trigonelline, an abundant xenobiotic alkaloid in coffee beans(26), and 1-methyl-2-pyridone-5-carboxamide, 3-methyl catechol sulfate, and 4-methyl catechol sulfate (27)), were significantly, positively associated with fibrinogen. Additionally, dimethylguanidino valerate (DMGV), a metabolite negatively correlated with fruit/vegetable consumption and self-reported physical activity (28), was positively associated with fibrinogen. Together, these compounds shed light on exogenous factors that may influence fibrinogen, inflammation, and coagulation.

### Metabolite Overlap with Prior Cardiovascular Disease Metabolomics Studies

To assess which metabolites associated with fibrinogen levels in our study agree with existing athero- thrombotic disease-metabolite associations and may therefore be likely to play a role in cardiovascular disease development (CVD), we reviewed 23 published, cohort-based large-scale metabolomics studies investigating plasma metabolite associations with 10 incident cardiovascular disease endpoints [CVD, overall cardiovascular mortality, venous thromboembolism (VTE), pulmonary embolism (PE), ischemic stroke (IS), overall incident stoke, coronary artery disease (CAD), coronary heart disease (CHD), myocardial infarction (MI), and heart failure (HF)] (see **Methods** for additional details). In total, we compiled a list of 297 plasma metabolites, corresponding to 429 metabolite-disease pairs, that have previously been significantly associated with these thrombotic disease endpoints. Of these metabolites, 171 were tested for association with fibrinogen in the present meta-analysis. 79 (∼46%) of these CVD-associated metabolites were also significantly associated with fibrinogen (**Supplemental Table 4**), of which the majority (N=58) were associated with concordant directions of effect (i.e. higher levels of the metabolite associated with higher vascular risk and higher fibrinogen, or vice versa). Histidine, beta-hydroxybutyrate, and phenylalanine were among fibrinogen-associated metabolites associating with a CVD outcome the greatest number of times in current literature.

## Discussion

Here we present a large-scale, untargeted metabolomics meta-analysis for circulating fibrinogen levels in a diverse population. The findings both validate and extend existing plasma metabolite-fibrinogen associations and implicate several metabolites which may reflect or drive alterations in thrombosis and inflammation, as well as downstream vascular conditions.

### Fibrinogen-associated peptides with links to vascular pathways

Positive associations observed between fibrinogen and peptide products of the complement and coagulation pathways validate the capture of relevant associations. Similar to prior studies(19), we observed several peptide fragments with potential links to the angiotensin and bradykinin peptide pathways, which increase IL-6(29) and when dysregulated, can result in phenotypes contributing to downstream cardiovascular disease (CVD) risk such as hypertension(30,31). Several fibrinogen-associated dipeptides are known cleavage products of ACE, including phenylalanylarginine, a direct product of bradykinin cleavage(32), and phenylalanylleucine(32). Additionally, circulating levels of phenylalanylserine, asparagylleucine, and alpha-glutamyl tyrosine have previously been associated with genetic variants in *ACE*, and are reported to be altered in ACE-inhibitor users(32). Hence, it is possible that these peptides reflect mechanisms connecting inflammation, coagulation, hypertension, and cardiovascular disease, but further studies are needed to investigate the sources of these dipeptides, and whether they play an active role in regulating vascular spasm and/or coagulation.

### Alterations in inflammatory lipids

Dyslipidemia is a known vascular risk factor(30), characterized by increased levels of circulating LDL-cholesterol, triglycerides, and free fatty acids, which are drivers of inflammation and atherosclerosis(33). Cholesterol and numerous triacylglycerol, diacylglycerol, and fatty-acyl species were positively associated with fibrinogen, and have previously been associated with CVD outcomes, indicating dyslipidemia is likely present in both disease and the underlying pro-coagulant and inflammatory risk state. Omega-6 fatty acids arachidonate (20:4n6) and dihomo-linolenate, as well as omega-3 fatty acids eicosapentaenoate (EPA; 20:5n3) and docosahexaenoate (DHA; 22:6n3) are eicosanoid precursors which mediate anti-inflammatory signaling(34), and were all inversely associated with fibrinogen. Similar to prior studies, we observed an enrichment for inverse associations between plasma membrane-constituent glycerophospholipid species and fibrinogen(20))(19), while sphingolipids were primarily positively associated with fibrinogen. Interestingly, a recent Mendelian Randomization study found evidence linking 4 lipids with risk for pro-hemorrhagic disorders; we observed similar derivatives of these lipids - (1- docosapentaenoylglycerophosphocholine (22:5n3), 1-palmitoylglycerophosphocholine (16:0), 1- docosahexaenoylglycerophosphocholine (22:6n3), palmitoyleoyl sphingomyelin) - significantly associated with fibrinogen levels, with opposite directions of effect on circulating fibrinogen as opposed to coagulation defects (35). While sensitivity analyses for C-Reactive Protein (CRP) suggest that a portion (∼6%) of metabolite associations may be heavily driven by inflammatory functions of fibrinogen, we note that this is only an approximate evaluation. Not all inflammatory pathways are captured by CRP, and confounding, such as differences in the accuracy of measures for fibrinogen and CRP, could contribute to incomplete adjustment for influences of inflammation.

We further acknowledge that the underlying source of pro-inflammatory metabolite alterations remains unclear. Interestingly, one fibrinogen gene beta (*FGB*) rare variant, rs6054 (allele frequency 0.004), which was robustly associated with fibrinogen levels in prior genome-wide association studies (GWAS)(9), also associates with lipid levels in other GWAS(36), implicating that some lipid- fibrinogen relationships may have genetic origins. However, environmental influences such as smoking can also drive inflammation (37). Further studies will be needed to investigate causal pathways and determine the direction of the relationship between genetic variants, lipids, and fibrinogen levels. Together, these associations may reflect a state of increased membrane damage in a procoagulant, proinflammatory state, although further work is needed to clarify the relationship between lipids and fibrinogen.

### Metabolite links between diet, coagulation, and inflammation

Several exogenous compounds were associated with fibrinogen and may indicate links between modifiable risk factors - such as diet and lifestyle - and vascular changes. In line with work establishing smoking as a CVD-risk factor (30) and known to promote a prothrombotic state(38), tobacco metabolite cotinine was positively associated with fibrinogen in this study, and has also been associated with fibrinogen and CRP in a prior study(19), demonstrating our ability to detect and validate epidemiological links in molecular data. Choline and betaine are both diet-derived compounds that are metabolized to trimethylamine N-oxide (TMAO) by gut microbiota(39–42) and were all associated with fibrinogen, adding to a body of literature investigating this pathway’s involvement in CVD pathogenesis(39–41).

Dimethylguanidino valerate (DMGV) was positively associated with fibrinogen and has positively been associated with a number of CVD outcomes(18,28,43,44) and risk factors(28,45), and inversely associated with healthy lifestyle factors, including dietary intake of vegetables and physical activity(28,45). Furthermore, the positive association observed between fibrinogen and caffeine supports previous small-scale clinical assessments, *in-vitro* assays, and animal studies which suggest that caffeine increases circulating coagulation factors(46,47). Together, these associations suggest potential markers of diet, lifestyle, and microbiome activity in proinflammatory or procoagulant state of CVD risk.

### Putative links between hormones and vascular pathways

Hormones and signaling molecules regulate metabolism and orchestrate responses to changing conditions. Here, we observed 3 amino-acid derived, and 8 cholesterol-derived, sex hormones associated with fibrinogen. Serotonin plays an active role in regulating vascular spasm during coagulation(48,49).

Cortisol and thyroid-T4 hormone thyroxine have suggestive links to CVD and/or coagulation. Cortisol has previously been associated with ischemic stroke(50), and higher levels of coagulation factors are reported in patients suffering from Cushing’s syndrome(51) or hyperthyroidism(52). However, other hormones, such as GABA have very little existing evidence for a role in coagulation, but are known to regulate ion homeostasis and activate diverse signaling patterns that could feasibly be related(53). Here, associations observed between fibrinogen and epiandrosterone, testosterone derivatives (5a-androstan- 3alpha,17alpha-diol (3a5a-adiol or 3a-diol), 5a-androstan-3alpha,17beta-diol monosulfate), and progesterone derivative 5-a pregnane-3(a or B), 20B diol disulfate, further support a postulated link between sex-hormone metabolism and coagulation, although none of these metabolites showed evidence of effect size heterogeneity between sexes.

### Metabolites with links to cardiovascular disease

In the present study, we observe associations with several metabolites that have previously been linked to CVD outcomes in large scale metabolomics studies. For example, several free amino acids such as histidine, tyrosine, phenylalanine, tryptophan, and alanine, and lipid species involved in inflammation, such as lysophosphocholines (LPCs), sphingomyelins (SMs), and triacylglycerols (TAGs) were significantly associated with cardiovascular disease outcomes in prior studies(54–59), and also associated with fibrinogen. Directions of associations were generally consistent between fibrinogen and CVD outcomes – with, for example, inverse associations between histidine levels and fibrinogen, as well as CVD outcomes of stroke(59), myocardial infarction(60), coronary artery disease(57), and coronary heart disease(61) reported in literature.

Not all metabolites previously associated with a CVD outcome were also associated with fibrinogen in the present study. It is possible that these metabolites impact CVD pathology through pathways independent of those captured by a fibrinogen measure. However, there are often substantial technical or sampling differences between metabolomics studies (for example sample size, demographics, preparation of plasma/serum samples, metabolomics technology employed to obtain measures, or analyses used to map spectra to metabolites) that can impact metabolite measures, and thereby influence significance of the associations observed. Furthermore, these differences may contribute to inconsistencies in effect directions among significant metabolites across studies. For example, not all effect-directions for a given metabolite and its disease associations were concordant in literature (with one study reporting a positive association between histidine and coronary heart disease(56), compared to negative associations observed in all other studies of CVD outcomes (59)(60)(57)(61)(43)). These discrepancies highlight the need for additional studies and harmonization of metabolite measures across platforms.

However, we captured several fibrinogen-associated metabolites that have not yet been associated with disease outcomes. We suggest some of these may reflect metabolites that influence the processes involved in pathogenesis of vascular diseases, which metabolomic studies associating metabolites with case-control disease endpoints may not yet be powered to detect significant associations with.

### Strengths and Limitations

The work presented here is, to our knowledge, among the first and largest efforts to assess metabolite profiles associated with a coagulation factor. Furthermore, the present study is more broadly representative of diverse populations within the US, in contrast to the few existing studies. Our study design enabled detection of many newly significant metabolite associations with fibrinogen which may reflect metabolites interacting with coagulation and inflammation to contribute to vascular disease risk. Despite these strengths, we note several limitations. First, we note that a fixed effects meta-analysis model does not account for potential relatedness across cohorts; specifically, FHS sub-studies may have familial correlation. However, no significant metabolites were based on FHS results alone. We also note that some of the CVD-metabolomics studies we compared metabolite associations from in literature review include data from the same cohorts assessed in the present study. Additionally, lack of universal ID reporting across metabolomics analyses creates challenges for consistently contextualizing metabolites in broader literature. For instance, the same metabolite can be reported with different nomenclature depending on the year, as is the case for 1-methylhistidine / 3-methylhistidine, and many studies do not report results with any standard nomenclature system (such as HMDB or RefMet IDs), making comparison of results across traits and across studies challenging. In this study, we mapped as many metabolites as possible to their associated HMDB ID’s to help create a consistent reference for future studies. Finally, the results presented are cross-sectional and do not provide information on causation or direction of effect. Future studies associating metabolites with additional quantitative factors, as well as functional studies, will be necessary to disentangle which metabolites causally influence or directly interact with pathways of coagulation and inflammation and may alter risk of downstream vascular disease.

## Methods

### Study design

Our study was comprised of cross-sectional data collected from six major U.S. community-based cohort studies including the Atherosclerosis Risk in Communities (ARIC) study(62), Cardiovascular Health Study (CHS)(63), Framingham Heart Study Offspring Cohort(64) and Third Generation (FHS)(65), Jackson Heart Study (JHS)(66), and Multi-Ethnic Study of Atherosclerosis (MESA)(67). Detailed descriptions of each study design were published previously and are briefly described in the **Supplemental Methods**. Each study was approved by Institutional Review Boards (IRB) at all participating institutions, and study participants provided written informed consent at all study visits.

This study was limited to participants with available information on fibrinogen levels, metabolite measures and covariates, at the same visit for all studies except CHS. The overlapping participants from ARIC and JHS (n=384) were excluded from ARIC. A total of 10,533 individuals were included in our analyses.

## Measurements

### Fibrinogen Measures

Fibrinogen concentration was measured in fasted citrated or EDTA plasma samples using a variety of methods including the Clauss method for ARIC, CHS, FHS, and JHS and immunonephelometric methods for MESA and FHS. Fibrinogen concentration was collected at the same exam as the metabolites for all studies except CHS. In CHS, fibrinogen was measured at baseline and 3 years later; the metabolome was measured 2 years after that.

### Metabolomic Measures

Metabolomic profiling was completed with measures by the Broad Institute (Boston, MA, USA) for MESA, CHS, JHS, and FHS and by Metabolon Inc. (Morrisville, NC, USA) for ARIC. Both platforms utilized gas chromatography–mass spectrometry (GC-MS) and/or liquid chromatography–mass spectrometry (LC-MS), and detailed protocols have been described elsewhere(18,68–70). In the current analysis, known metabolites with missing rates <25% within a contributing study were included. Within each study, metabolite levels were first winsorized at ± 4 standard deviations. Missing metabolite values were then imputed using ½ the minimum value, as recommended by recent work in TOPMed (71); and metabolite values were standardized with mean=0 and SD=1 prior to the analysis. We considered 1,282 unique metabolites from the individual studies for the meta-analysis. After filtering for metabolites present in 2 or more cohorts, with missingness =< 25%, 789 metabolites were tested in the meta-analysis.

### Covariates of Interest

Covariates were measured at the same exam as the metabolites for all studies. Covariates included age (years), sex, BMI (kg/m^2^), smoking status (current/former/never), alcohol consumption (current/former/never), low-density lipoprotein (LDL), high-density lipoprotein (HDL), triglycerides, diabetes status, and study specific variables such as center and batch based on known impacts of these covariates on fibrinogen variance. For all studies, HDL-C was directly assayed, while LDL-C was estimated using the Friedewald equation(72). CHS was further adjusted for the time-lag to reflect the time difference between metabolite and fibrinogen collection. To evaluate the impact of this time lapse, we performed sensitivity analyses excluding CHS and found that nearly all (257/270) metabolite associations remained significant (**Supplemental Table 14**).

## Statistical Analysis

Linear regression was used to assess the relationship between each metabolite and fibrinogen by study and self-reported race, as a rough proxy for a variety of social and environmental influences which are often unmeasured or inconsistently measured across cohorts. For cohorts with family structures, linear mixed models were applied. First, each study calculated the fibrinogen residuals adjusting for age, sex, BMI, smoking status, drinking status, LDL, HDL, triglycerides, diabetes status, and study specific variables (such as batch and center). Second, inverse normal transformed fibrinogen residuals were calculated. Third, the inverse normal transformed residuals were scaled by the standard deviation of the fibrinogen residuals calculated in step one to better reflect the original fibrinogen distributions within each cohort. The rescaled inverse normal transformed fibrinogen levels were used for analyses with metabolite levels, adjusting for the same covariates previously described. Meta-analysis was performed using the META package in R. We obtained pooled effect size estimates and standard deviations using inverse variance weighting for each metabolite. Metabolites with pooled *p*-values below FDR at 0.05 were considered statistically significant.

To evaluate which metabolite associations may be primarily driven by inflammation or kidney function, we conducted sensitivity analyses for C-reactive protein (CRP) in JHS (n=2,250) and Estimated Glomerular Filtration Rate (eGFR) in ARIC (n=1,929), respectively, as these were the studies with available concurrent data. We considered a p-value fold-change of at least 2 towards non-significance as evidence of attenuation of the association following adjustment for the additional covariate. We also performed sex-stratified analyses to estimate potential sex-specific effects. Effect-size heterogeneity between male and female estimated via Cochran’s Q and I^2^ tests.

### Metabolite Classification

Metabolite names were mapped to standard Human Metabolome Database (HMDB) and Kyoto Encyclopedia of Genes and Genomes (KEGG) identifiers to facilitate standardized classification of metabolites and reproducible analysis between this study and future metabolomics studies. The HMDB Metabolome 5.0 (https://hmdb.ca/downloads, downloaded 3/30/2022) was used to classify all metabolites tested in the meta-analysis by superclass, class, subclass, and direct parent by HMDB ID. Eighty-nine metabolites that did not automatically map to HMDB IDs in study submitted data (for example, many lipid related molecules) were directly queried in the HMDB and manually matched to an HMDB ID if a synonymous metabolite name with an associated ID could be found. For metabolites with matched HMDB IDs, HMDB superclass, class, subclass, and parent categories were reported. Metabolites without HMDB IDs were manually classified into these categories by HMDB criteria where possible.

### Enrichment and Pathway Analysis

Each HMDB superclass, class, sub-class, and direct-parent category was tested for enrichment in significant metabolites using Fisher’s Exact Tests, which compared the counts of tested metabolites in each category to the counts of metabolites in each class that were significantly associated with fibrinogen following meta-analysis. Within each class, we used the binomial test to determine whether there was statistical enrichment for positive vs. inverse associations. MetaboAnalyst 5.0 was used to assess Small Molecular Pathway Database (SMPDB) and KEGG pathway enrichment for the 217/270 significantly associated metabolites that mapped to HMDB identifiers, as compared to the reference set of all tested metabolites. No pathways were statistically enriched in these analyses (FDR p>0.05 for all pathways), so results are not presented; however, as many metabolites did not map to existing identifiers within these databases (such as KEGG, for which 147 of 270 significant metabolites mapped), we consider this pathway analysis only preliminary. KEGG pathway maps were used to guide analysis of metabolites and interconnections between pathways in the results.

### Characterizing Observed Associations Relative to Previously Reported Inflammatory-Marker or Cardiovascular Disease Associations

To contextualize findings, we compared metabolites significantly associated with circulating fibrinogen here to metabolites previously associated with an inflammatory marker or a cardiovascular disease outcome in a study published prior to August 2023. Studies often report results corresponding to multiple covariate adjustment strategies and different significance thresholds. To maximize consistency, we used published results derived from models with minimal covariate adjustment and the least stringent reported p-value significance threshold to define significant associations. To determine metabolites associated with inflammatory markers, we used significant results from a 2017 study of 324 serum metabolite associations with circulating interleukin-6 (IL-6) in 73 elderly adults(21), a 2022 study of 860 plasma metabolites and their associations with circulating C-Reactive protein (CRP) and fibrinogen in 1,234 patients with coronary artery disease(20), and a 2017 study of 613 plasma metabolite associations with circulating fibrinogen, CRP, and white blood cell count in 925 healthy adults(19).

To determine metabolites associated with CVD, we compiled studies using untargeted metabolomics, or a platform targeting >=50 metabolites, with at least 400 participants in the discovery cohort. In total, we used 23 studies for 10 incident CVD endpoints [CVD, overall cardiovascular mortality, venous thromboembolism (VTE), pulmonary embolism (PE), ischemic stroke (IS), overall incident stoke, coronary artery disease (CAD), coronary heart disease (CHD), myocardial infarction (MI), and heart failure (HF)]. Studies are described in more detail in **Supplemental Table 13**.

## Data Availability

Summary level data for the meta-analysis and each contributing cohort is present in the supplement. Individual level data can be obtained through dbGap, BioLINCC or through individual cohort coordinating centers.

## Acknowledgments

This work was funded by R01HL139553. The project described was also supported by the National Center for Advancing Translational Sciences, National Institutes of Health, through Grant KL2TR002490 (LMR).

Molecular data for the Trans-Omics in Precision Medicine (TOPMed) program was supported by the National Heart, Lung and Blood Institute (NHLBI). Metabolomics for NHLBI TOPMed: MESA phs001416 was performed at Broad Metabolomics (HHSN268201600038I). Core support including phenotype harmonization, data management, sample-identity QC, and general program coordination were provided by the TOPMed Data Coordinating Center (R01HL-120393; U01HL-120393; contract HHSN268201800001I). We gratefully acknowledge the studies and participants who provided biological samples and data for TOPMed.

The Atherosclerosis Risk in Communities study has been funded in whole or in part with Federal funds from the National Heart, Lung, and Blood Institute, National Institutes of Health, Department of Health and Human Services, under Contract nos. (75N92022D00001, 75N92022D00002, 75N92022D00003, 75N92022D00004, 75N92022D00005). The authors thank the staff and participants of the ARIC study for their important contributions.

Cardiovascular Health Study: This research was supported by contracts HHSN268201200036C, HHSN268200800007C, HHSN268201800001C, N01HC55222, N01HC85079, N01HC85080, N01HC85081, N01HC85082, N01HC85083, N01HC85086, 75N92021D00006, and grants U01HL080295, HL105756, U01HL130114, R01HL172803, R01HL128575, R01HL087652, R01HL103612, R01HL105756, R01HL120393, from the National Heart, Lung, and Blood Institute (NHLBI), with additional contribution from the National Institute of Neurological Disorders and Stroke (NINDS). Additional support was provided by R01AG023629 from the National Institute on Aging (NIA). A full list of principal CHS investigators and institutions can be found at CHS-NHLBI.org. The content is solely the responsibility of the authors and does not necessarily represent the official views of the National Institutes of Health.

The Jackson Heart Study (JHS) is supported and conducted in collaboration with Jackson State University (HHSN268201800013I), Tougaloo College (HHSN268201800014I), the Mississippi State Department of Health (HHSN268201800015I) and the University of Mississippi Medical Center (HHSN268201800010I, HHSN268201800011I and HHSN268201800012I) contracts from the National Heart, Lung, and Blood Institute (NHLBI) and the National Institute on Minority Health and Health Disparities (NIMHD). The authors also wish to thank the staffs and participants of the JHS.

Multi-Ethnic Study of Atherosclerosis (MESA) (phs001416.v3.p1) was performed at the Broad Institute of MIT and Harvard (3U54HG003067-13S1). Centralized read mapping and genotype calling, along with variant quality metrics and filtering were provided by the TOPMed Informatics Research Center (3R01HL-117626-02S1). Phenotype harmonization, data management, sample-identity QC, and general study coordination, were provided by the TOPMed Data Coordinating Center (3R01HL-120393-02S1), and TOPMed MESA Multi-Omics (HHSN2682015000031/HSN26800004). The MESA projects are conducted and supported by the National Heart, Lung, and Blood Institute (NHLBI) in collaboration with MESA investigators. Support for the Multi-Ethnic Study of Atherosclerosis (MESA) projects are conducted and supported by the National Heart, Lung, and Blood Institute (NHLBI) in collaboration with MESA investigators. Support for MESA is provided by contracts 75N92020D00001, HHSN268201500003I, N01-HC-95159, 75N92020D00005, N01-HC-95160, 75N92020D00002, N01-HC-95161, 75N92020D00003, N01-HC-95162, 75N92020D00006, N01-HC-95163, 75N92020D00004, N01-HC-95164, 75N92020D00007, N01-HC-95165, N01-HC-95166, N01-HC-95167, N01-HC-95168, N01-HC-95169, UL1-TR-000040, UL1-TR-001079, UL1-TR-001420, UL1TR001881, DK063491, and R01HL105756. The authors thank the other investigators, the staff, and the participants of the MESA study for their valuable contributions. A full list of participating MESA investigators and institutes can be found at http://www.mesa-nhlbi.org. Also supported in part by the National Heart, Lung, and Blood Institute (NHLBI) R01HL168683.

The views expressed in this manuscript are those of the authors and do not necessarily represent the views of the National Heart, Lung, and Blood Institute; the National Institutes of Health; or the U.S. Department of Health and Human Services.

Cartoon images created in Biorender (www.biorender.com).

---

**Supplemental Figure 1.**
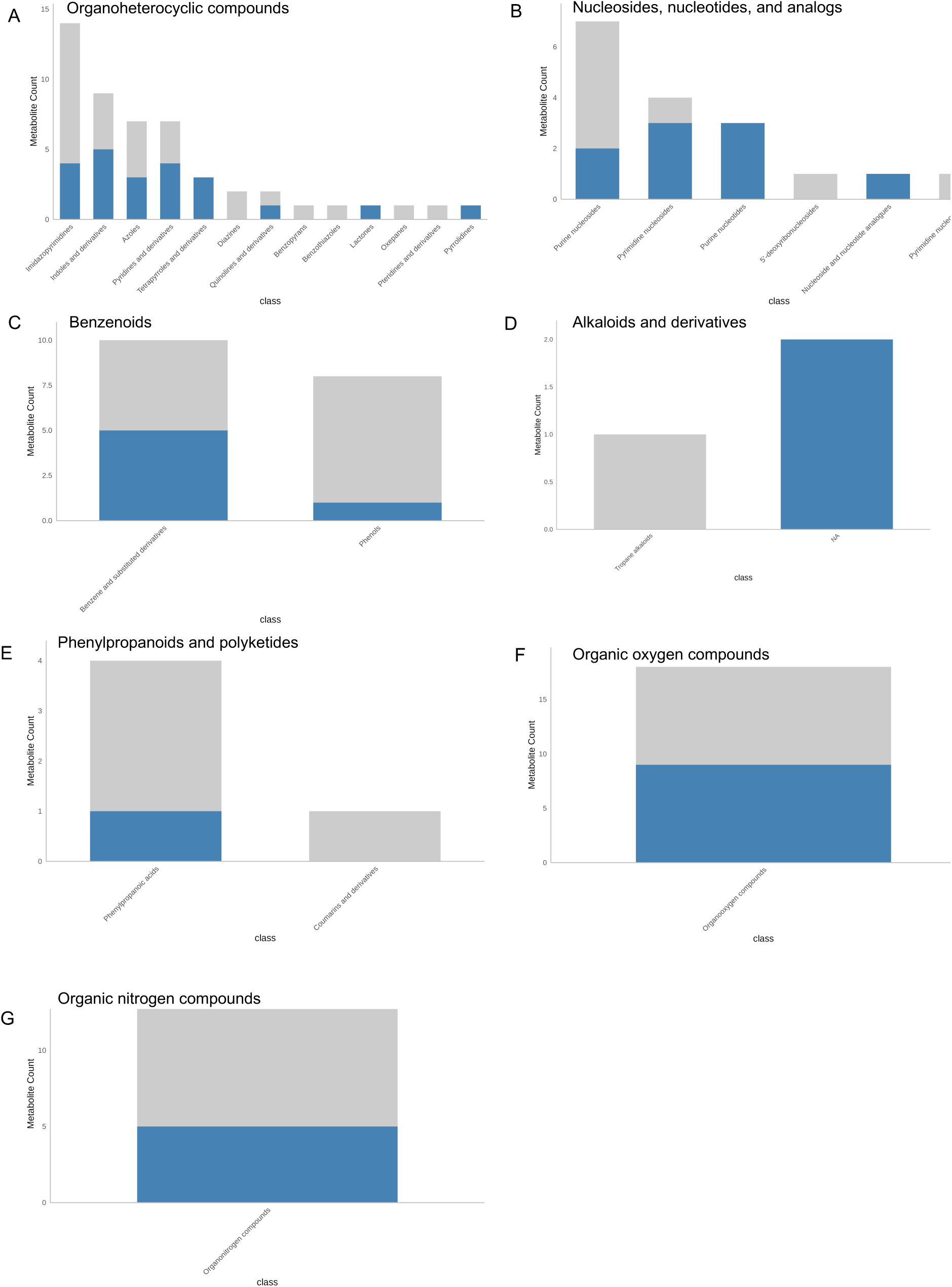
Metabolites within each HMDB class tested (gray) and significant (blue) in the meta-analysis, with each plot showing metabolites within a different HMDB super-class. (A) Organoheterocyclic compounds. (B) Nucleosides, nucleotides, and analogs. (C) Benzenoids. (D) Alkaloids and derivatives. (E) Phenylpropanoids and polyketides. (F) Organic oxygen compounds. (G) Organic nitrogen compounds.

**Supplemental Figure 2.**
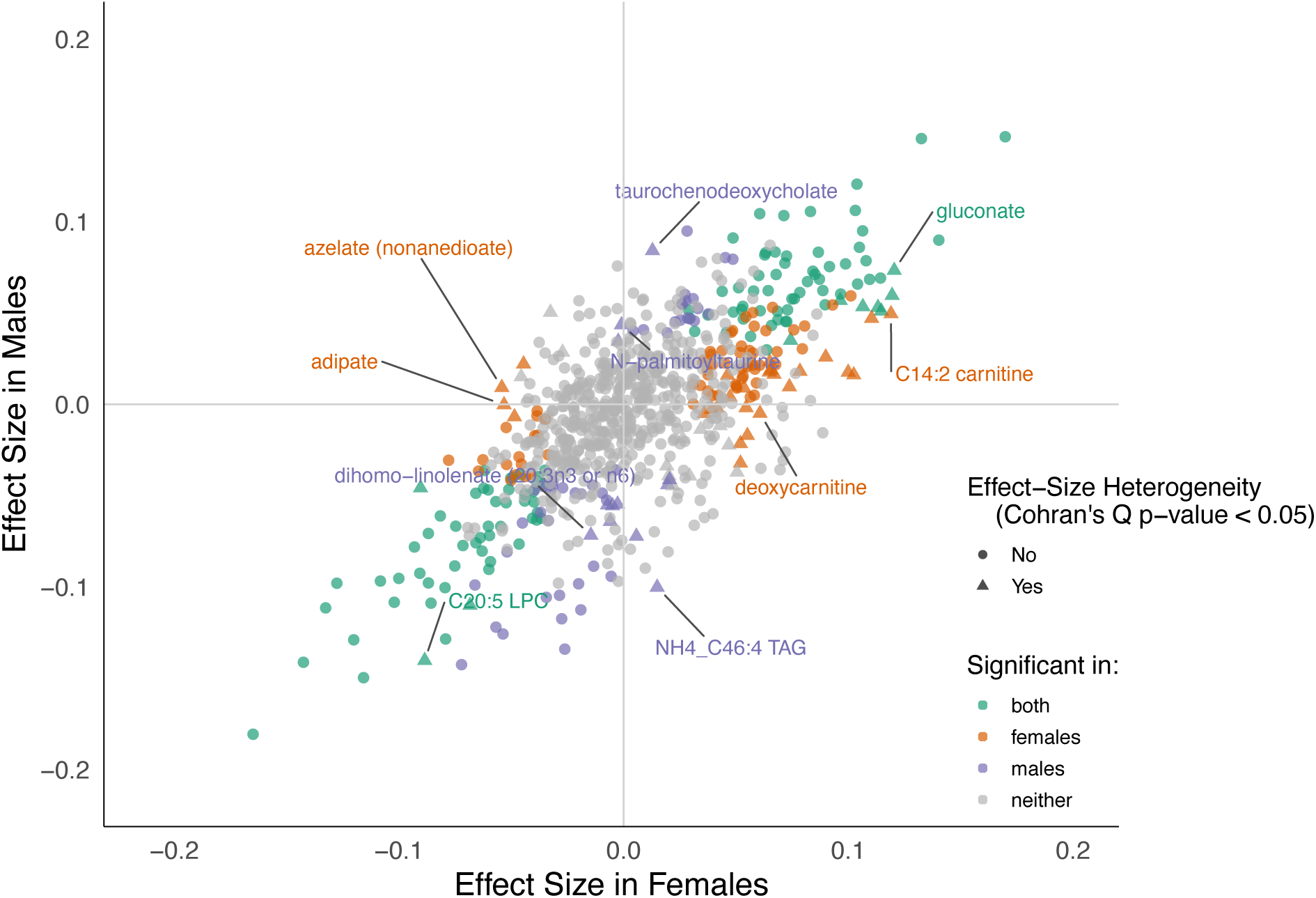
Metabolites tested in the sex-combined and sex-stratified meta-analyses, according to whether significant in both male and female sex-stratified analyses (green), female sex-stratified analysis (orange), male sex-stratified analysis (purple), or neither sex-stratified analysis. Shape of points depicts whether effect-size heterogeneity was observed between sexes (Cochran’s Q p-value<0.05).

**Supplemental Figure 3.**
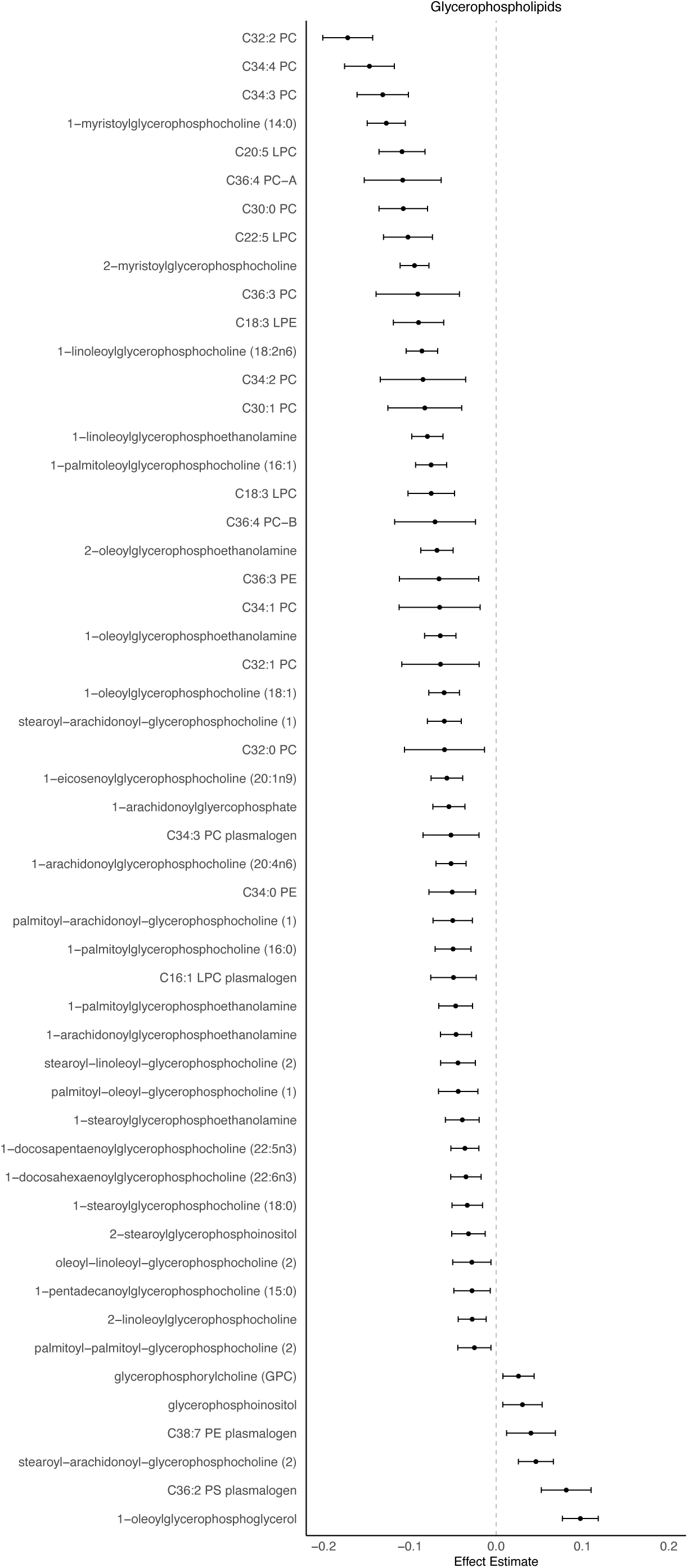
Forest plot depicting meta-analysis effect-sizes and standard errors for glycerophospholipids significantly associated with fibrinogen. Glycerophospholipids are significantly enriched for negative effect-size associations with fibrinogen (47/53 metabolite effects negative, binomial test FDR-adjusted p-value = 1.74 x 10^-7^).

**Supplemental Figure 4.**
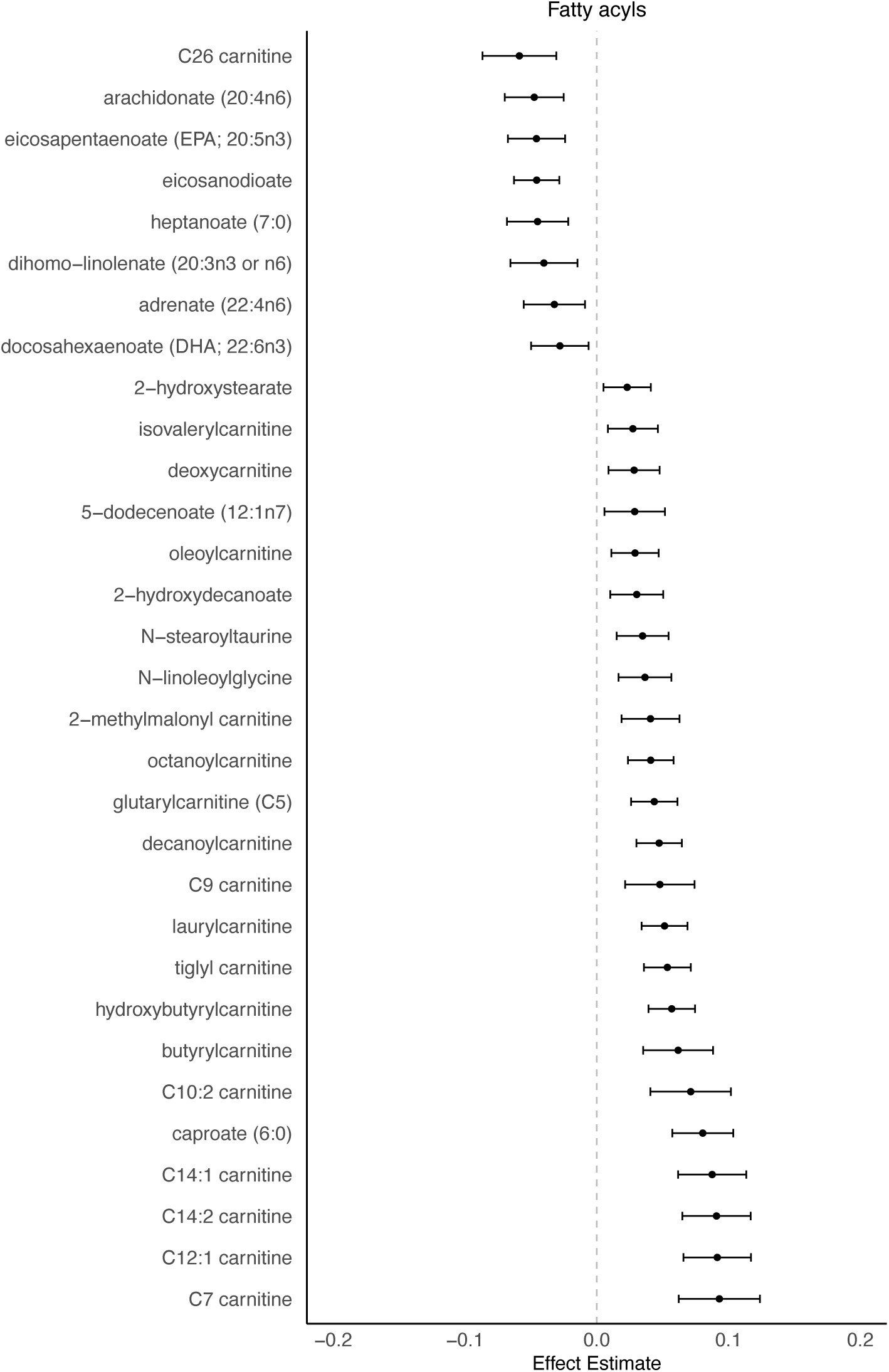
Forest plot depicting meta-analysis effect-sizes and standard errors for fatty acyls significantly associated with fibrinogen. Fatty acyls were nominally enriched for positive associations (23/31 associations positive, binomial p-value = 0.02, FDR-adjusted p-value = 0.11).

**Supplemental Figure 5.**
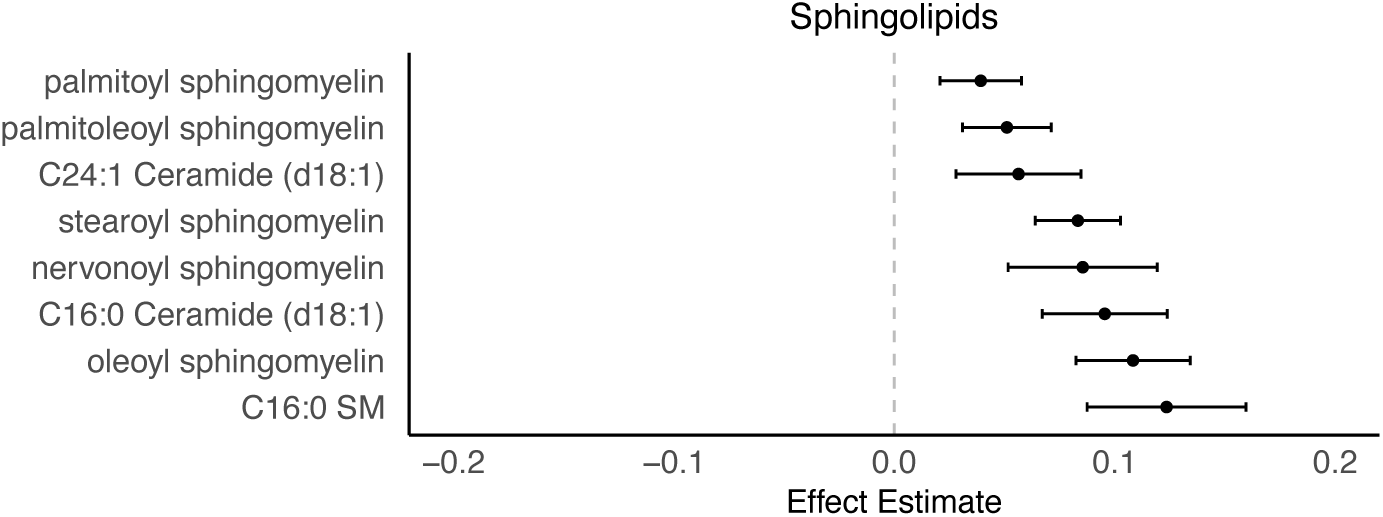
Forest plot depicting meta-analysis effect-sizes and standard errors for sphingolipids significantly associated with fibrinogen. Sphingolipids were nominally enriched for positive associations (8/8 associations positive, binomial p-value = 0.007, FDR-adjusted p-value = 0.11).

**Supplemental Figure 6.**
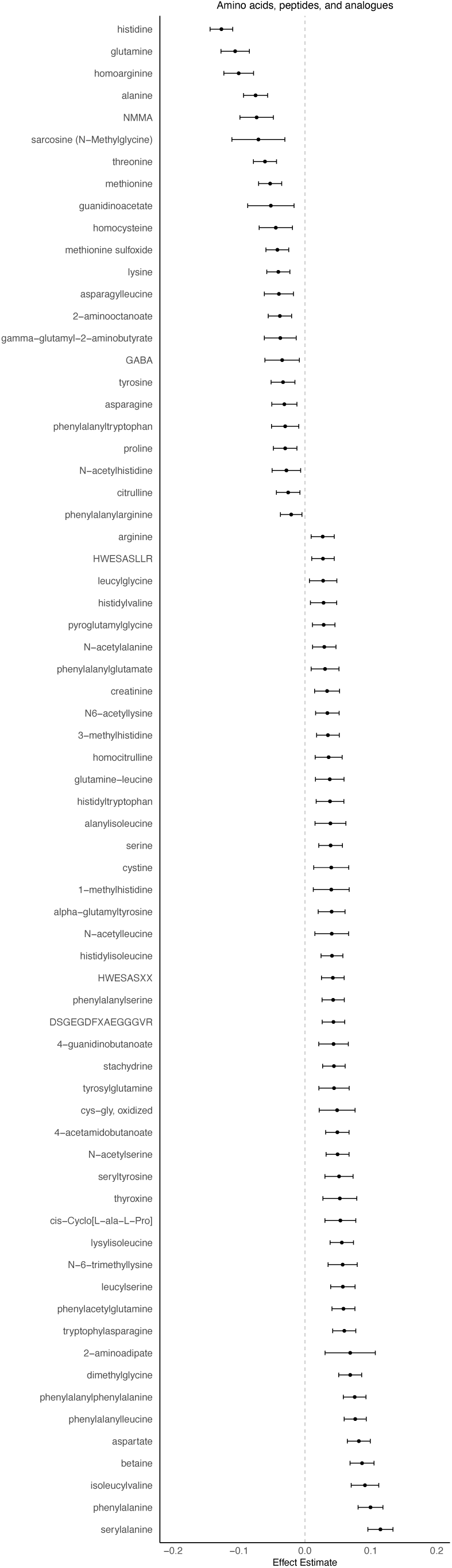
Forest plot depicting meta-analysis effect-sizes and standard errors for amino acids and peptides significantly associated with fibrinogen.

**Supplemental Figure 7.**
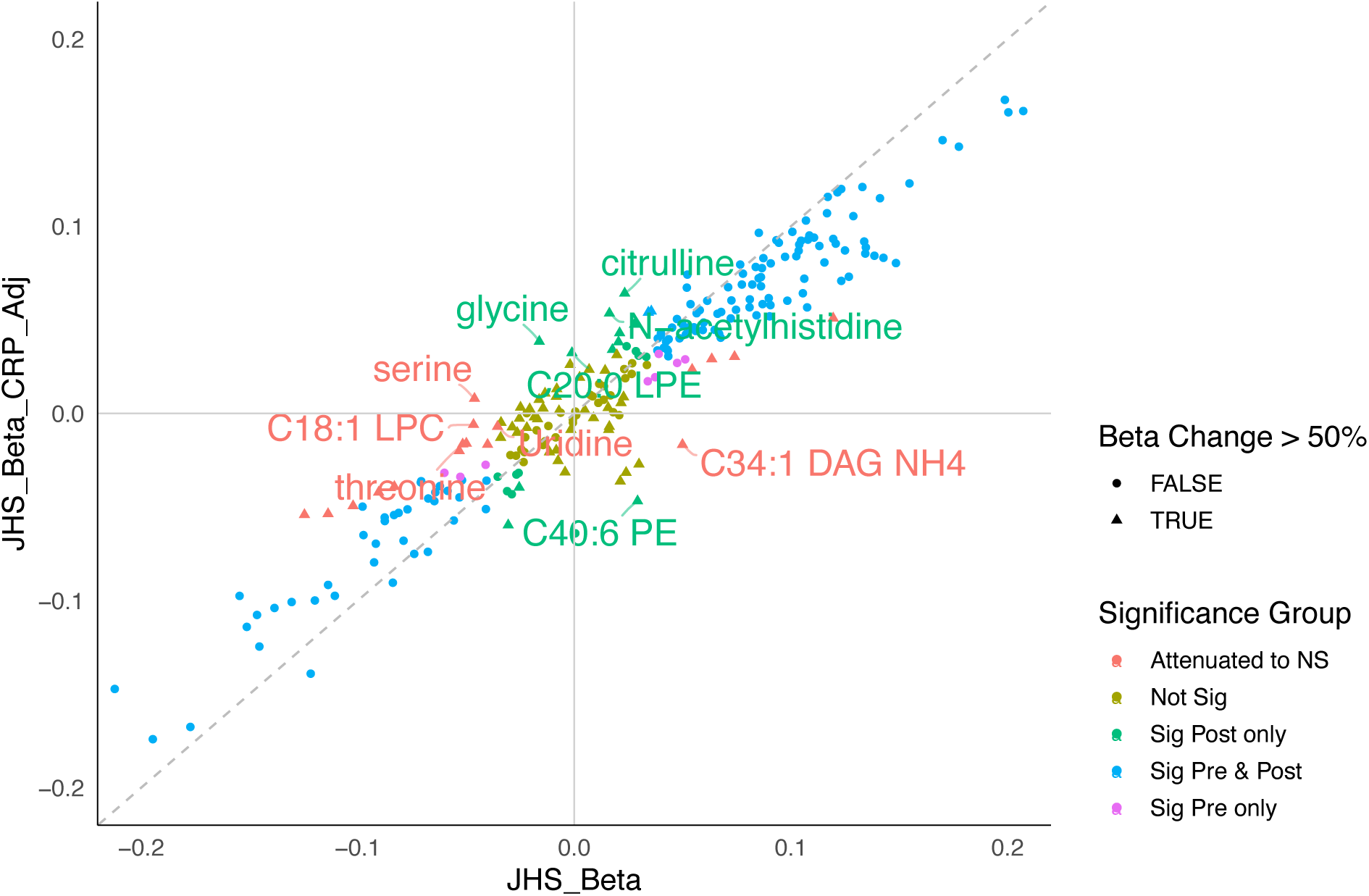
Jackson Heart Study (JHS) effect-sizes for metabolite association with fibrinogen in standard covariate model and C-Reactive Protein (CRP)-adjusted model, with points colored according to whether significant in model not adjusted for CRP, model adjusted for CRP, or neither model, and shaped according to whether the effect size changed by at least 50% between models.

## References

1. Wolberg AS, Sang Y. Fibrinogen and factor XIII in venous thrombosis and thrombus stability. Arterioscler Thromb Vasc Biol. 2022 Aug;42(8):931–41.

2. Sabater-Lleal M, Huang J, Chasman D, Naitza S, Dehghan A, Johnson AD, et al. Multiethnic meta-analysis of genome-wide association studies in >100 000 subjects identifies 23 fibrinogen-associated Loci but no strong evidence of a causal association between circulating fibrinogen and cardiovascular disease. Circulation. 2013 Sep 17;128(12):1310–24.

3. Ward-Caviness CK, de Vries PS, Wiggins KL, Huffman JE, Yanek LR, Bielak LF, et al. Mendelian randomization evaluation of causal effects of fibrinogen on incident coronary heart disease. PLoS ONE. 2019 May 10;14(5):e0216222.

4. Vilar R, Fish RJ, Casini A, Neerman-Arbez M. Fibrin(ogen) in human disease: both friend and foe. Haematologica. 2020 Jan 31;105(2):284–96.

5. Aleman MM, Walton BL, Byrnes JR, Wolberg AS. Fibrinogen and red blood cells in venous thrombosis. Thromb Res. 2014 May;133 Suppl 1(0 1):S38-40.

6. Danesh J, Lewington S, Thompson SG, Lowe GDO, Collins R, Kostis JB, et al. Plasma fibrinogen level and the risk of major cardiovascular diseases and nonvascular mortality: an individual participant meta-analysis. JAMA. 2005 Oct 12;294(14):1799–809.

7. Lutsey PL, Cushman M, Steffen LM, Green D, Barr RG, Herrington D, et al. Plasma hemostatic factors and endothelial markers in four racial/ethnic groups: the MESA study. J Thromb Haemost. 2006 Dec;4(12):2629–35.

8. Bogar L, Juricskay I, Kesmarky G, Feher G, Kenyeres P, Toth K. Gender differences in hemorheological parameters of coronary artery disease patients. Clin Hemorheol Microcirc. 2006;35(1–2):99–103.

9. Huffman JE, Nicholas J, Hahn J, Heath AS, Raffield LM, Yanek LR, et al. Whole-genome analysis of plasma fibrinogen reveals population-differentiated genetic regulators with putative liver roles. Blood. 2024 Nov 21;144(21):2248–65.

10. de Vries PS, Chasman DI, Sabater-Lleal M, Chen M-H, Huffman JE, Steri M, et al. A meta-analysis of 120 246 individuals identifies 18 new loci for fibrinogen concentration. Hum Mol Genet. 2016 Jan 15;25(2):358–70.

11. de Vries PS, Sabater-Lleal M, Chasman DI, Trompet S, Ahluwalia TS, Teumer A, et al. Comparison of HapMap and 1000 Genomes Reference Panels in a Large-Scale Genome-Wide Association Study. PLoS ONE. 2017 Jan 20;12(1):e0167742.

12. Huffman JE, de Vries PS, Morrison AC, Sabater-Lleal M, Kacprowski T, Auer PL, et al. Rare and low-frequency variants and their association with plasma levels of fibrinogen, FVII, FVIII, and vWF. Blood. 2015 Sep 10;126(11):e19–29.

13. Pankratz N, Wei P, Brody JA, Chen M-H, de Vries PS, Huffman JE, et al. Whole-exome sequencing of 14 389 individuals from the ESP and CHARGE consortia identifies novel rare variation associated with hemostatic factors. Hum Mol Genet. 2022 Sep 10;31(18):3120–32.

14. Balleisen L, Bailey J, Epping PH, Schulte H, van de Loo J. Epidemiological study on factor VII, factor VIII and fibrinogen in an industrial population: I. Baseline data on the relation to age, gender, body-weight, smoking, alcohol, pill-using, and menopause. Thromb Haemost. 1985 Aug 30;54(2):475–9.

15. D’Alessandro AJ. Vitamin K and its rôle in blood coagulation. The American Journal of Surgery. 1942 Jul;57(1):104–11.

16. Moncada S, Vane JR. Prostacyclin and blood coagulation. Drugs. 1981 Jun;21(6):430–7.

17. Jung E, Kong SY, Ro YS, Ryu HH, Shin SD. Serum Cholesterol Levels and Risk of Cardiovascular Death: A Systematic Review and a Dose-Response Meta-Analysis of Prospective Cohort Studies. Int J Environ Res Public Health. 2022 Jul 6;19(14).

18. Tahir UA, Katz DH, Zhao T, Ngo D, Cruz DE, Robbins JM, et al. Metabolomic profiles and heart failure risk in black adults: insights from the jackson heart study. Circ Heart Fail. 2021 Jan 19;14(1):e007275.

19. Pietzner M, Kaul A, Henning A-K, Kastenmüller G, Artati A, Lerch MM, et al. Comprehensive metabolic profiling of chronic low-grade inflammation among generally healthy individuals. BMC Med. 2017 Nov 30;15(1):210.

20. Zhu Q, Wu Y, Mai J, Guo G, Meng J, Fang X, et al. Comprehensive metabolic profiling of inflammation indicated key roles of glycerophospholipid and arginine metabolism in coronary artery disease. Front Immunol. 2022 Mar 8;13:829425.

21. Lustgarten MS, Fielding RA. Metabolites Associated With Circulating Interleukin-6 in Older Adults. J Gerontol A Biol Sci Med Sci. 2017 Sep 1;72(9):1277–83.

22. Hahn J, Temprano-Sagrera G, Hasbani NR, Ligthart S, Dehghan A, Wolberg AS, et al. Bivariate genome-wide association study of circulating fibrinogen and C-reactive protein levels. J Thromb Haemost. 2024 Dec;22(12):3448–59.

23. Wang H, Zheng C, Lu Y, Jiang Q, Yin R, Zhu P, et al. Urinary fibrinogen as a predictor of progression of CKD. Clin J Am Soc Nephrol. 2017 Dec 7;12(12):1922–9.

24. Liese AD, Weis KE, Schulz M, Tooze JA. Food intake patterns associated with incident type 2 diabetes: the Insulin Resistance Atherosclerosis Study. Diabetes Care. 2009 Feb;32(2):263–8.

25. Miura K, Nakagawa H, Ueshima H, Okayama A, Saitoh S, Curb JD, et al. Dietary factors related to higher plasma fibrinogen levels of Japanese-americans in hawaii compared with Japanese in Japan. Arterioscler Thromb Vasc Biol. 2006 Jul;26(7):1674–9.

26. Wu X, Skog K, Jägerstad M. Trigonelline, a naturally occurring constituent of green coffee beans behind the mutagenic activity of roasted coffee? Mutation Research/Genetic Toxicology and Environmental Mutagenesis. 1997 Jul;391(3):171–7.

27. He WJ, Chen J, Razavi AC, Hu EA, Grams ME, Yu B, et al. Metabolites Associated with Coffee Consumption and Incident Chronic Kidney Disease. Clin J Am Soc Nephrol. 2021 Nov 4;16(11):1620–9.

28. Ottosson F, Ericson U, Almgren P, Smith E, Brunkwall L, Hellstrand S, et al. Dimethylguanidino Valerate: A Lifestyle-Related Metabolite Associated With Future Coronary Artery Disease and Cardiovascular Mortality. J Am Heart Assoc. 2019 Oct;8(19):e012846.

29. Luther JM, Gainer JV, Murphey LJ, Yu C, Vaughan DE, Morrow JD, et al. Angiotensin II induces interleukin-6 in humans through a mineralocorticoid receptor-dependent mechanism. Hypertension. 2006 Dec;48(6):1050–7.

30. Garg R, Aggarwal S, Kumar R, Sharma G. Association of atherosclerosis with dyslipidemia and co-morbid conditions: A descriptive study. J Nat Sci Biol Med. 2015 Jun;6(1):163–8.

31. Herman LL, Padala SA, Ahmed I, Bashir K. Angiotensin-Converting Enzyme Inhibitors (ACEI). StatPearls. Treasure Island (FL): StatPearls Publishing; 2025.

32. Altmaier E, Menni C, Heier M, Meisinger C, Thorand B, Quell J, et al. The Pharmacogenetic Footprint of ACE Inhibition: A Population-Based Metabolomics Study. PLoS ONE. 2016 Apr 27;11(4):e0153163.

33. Klop B, Elte JWF, Cabezas MC. Dyslipidemia in obesity: mechanisms and potential targets. Nutrients. 2013 Apr 12;5(4):1218–40.

34. Yamaguchi A, Botta E, Holinstat M. Eicosanoids in inflammation in the blood and the vessel. Front Pharmacol. 2022 Sep 27;13:997403.

35. Wei J, Yang Z, Wu X, Zheng N, Wu D. Unveiling the role of lipid metabolism in haemorrhagic disorders: genetic insights and therapeutic perspectives. Thromb J. 2025 May 31;23(1):55.

36. Graham SE, Clarke SL, Wu K-HH, Kanoni S, Zajac GJM, Ramdas S, et al. The power of genetic diversity in genome-wide association studies of lipids. Nature. 2021 Dec 9;600(7890):675–9.

37. Elisia I, Lam V, Cho B, Hay M, Li MY, Yeung M, et al. The effect of smoking on chronic inflammation, immune function and blood cell composition. Sci Rep. 2020 Nov 10;10(1):19480.

38. Pretorius E, Oberholzer HM, van der Spuy WJ, Meiring JH. Smoking and coagulation: the sticky fibrin phenomenon. Ultrastruct Pathol. 2010 Aug;34(4):236–9.

39. Pan X-F, Yang JJ, Shu X-O, Moore SC, Palmer ND, Guasch-Ferré M, et al. Associations of circulating choline and its related metabolites with cardiometabolic biomarkers: an international pooled analysis. Am J Clin Nutr. 2021 Sep 1;114(3):893–906.

40. Medeiros GCBS de, Azevedo KPM de, Mesquita GXB, Lima SCVC, Silva DF de O, Pimenta IDSF, et al. Red meat consumption, risk of incidence of cardiovascular disease and cardiovascular mortality, and the dose-response effect: Protocol for a systematic review and meta-analysis of longitudinal cohort studies. Medicine (Baltimore). 2019 Sep;98(38):e17271.

41. Obeid R. The metabolic burden of methyl donor deficiency with focus on the betaine homocysteine methyltransferase pathway. Nutrients. 2013 Sep 9;5(9):3481–95.

42. Wang Z, Klipfell E, Bennett BJ, Koeth R, Levison BS, Dugar B, et al. Gut flora metabolism of phosphatidylcholine promotes cardiovascular disease. Nature. 2011 Apr 7;472(7341):57–63.

43. Cruz DE, Tahir UA, Hu J, Ngo D, Chen Z-Z, Robbins JM, et al. Metabolomic analysis of coronary heart disease in an african american cohort from the jackson heart study. JAMA Cardiol. 2022 Feb 1;7(2):184–94.

44. Ament Z, Patki A, Chaudhary N, Bhave VM, Garcia Guarniz A-L, Gao Y, et al. Nucleosides associated with incident ischemic stroke in the REGARDS and JHS cohorts. Neurology. 2022 May 24;98(21):e2097–107.

45. Wali JA, Koay YC, Chami J, Wood C, Corcilius L, Payne RJ, et al. Nutritional and metabolic regulation of the metabolite dimethylguanidino valeric acid: an early marker of cardiometabolic disease. Am J Physiol Endocrinol Metab. 2020 Sep 1;319(3):E509–18.

46. Nagelkirk PR, Sackett JR, Aiello JJ, Fitzgerald LF, Saunders MJ, Hargens TA, et al. Caffeine augments the prothrombotic but not the fibrinolytic response to exercise. Med Sci Sports Exerc. 2019 Mar;51(3):421–5.

47. Santhakumar A, Fozzard N, Perkins A, Singh I. Activity and Hemostatic Function.

48. Cloutier N, Allaeys I, Marcoux G, Machlus KR, Mailhot B, Zufferey A, et al. Platelets release pathogenic serotonin and return to circulation after immune complex-mediated sequestration. Proc Natl Acad Sci USA. 2018 Feb 13;115(7):E1550–9.

49. Van Nueten JM, Janssens WJ, Vanhoutte PM. Serotonin and vascular reactivity. Pharmacol Res Commun. 1985 Jul;17(7):585–608.

50. Balasubramanian R, Paynter NP, Giulianini F, Manson JE, Zhao Y, Chen J-C, et al. Metabolomic profiles associated with all-cause mortality in the Women’s Health Initiative. Int J Epidemiol. 2020 Feb 1;49(1):289–300.

51. Coelho MCA, Vieira Neto L, Kasuki L, Wildemberg LE, Santos CV dos, Castro G, et al. Rotation thromboelastometry and the hypercoagulable state in Cushing’s syndrome. Clin Endocrinol (Oxf). 2014 Nov;81(5):657–64.

52. Engelmann B, Bischof J, Dirk A-L, Friedrich N, Hammer E, Thiele T, et al. Effect of Experimental Thyrotoxicosis onto Blood Coagulation: A Proteomics Study. Eur Thyroid J. 2015 Sep;4(Suppl 1):119–24.

53. Ciranna L. Serotonin as a modulator of glutamate- and GABA-mediated neurotransmission: implications in physiological functions and in pathology. Curr Neuropharmacol. 2006 Apr;4(2):101–14.

54. McGranaghan P, Saxena A, Rubens M, Radenkovic J, Bach D, Schleußner L, et al. Predictive value of metabolomic biomarkers for cardiovascular disease risk: a systematic review and meta-analysis. Biomarkers. 2020 Mar;25(2):101–11.

55. Guo Y, Chen S-F, Zhang Y-R, Wang H-F, Huang S-Y, Chen S-D, et al. Circulating metabolites associated with incident myocardial infarction and stroke: A prospective cohort study of 90 438 participants. J Neurochem. 2022 Aug;162(4):371–84.

56. Cavus E, Karakas M, Ojeda FM, Kontto J, Veronesi G, Ferrario MM, et al. Association of circulating metabolites with risk of coronary heart disease in a european population: results from the biomarkers for cardiovascular risk assessment in europe (biomarcare) consortium. JAMA Cardiol. 2019 Dec 1;4(12):1270–9.

57. Ritchie SC, Surendran P, Karthikeyan S, Lambert SA, Bolton T, Pennells L, et al. Quality control and removal of technical variation of NMR metabolic biomarker data in ∼120,000 UK Biobank participants. Sci Data. 2023 Jan 31;10(1):64.

58. Würtz P, Havulinna AS, Soininen P, Tynkkynen T, Prieto-Merino D, Tillin T, et al. Metabolite profiling and cardiovascular event risk: a prospective study of 3 population-based cohorts. Circulation. 2015 Mar 3;131(9):774–85.

59. Vojinovic D, Kalaoja M, Trompet S, Fischer K, Shipley MJ, Li S, et al. Association of circulating metabolites in plasma or serum and risk of stroke: Meta-analysis from seven prospective cohorts. Neurology. 2020 Dec 2;96(8):e1110–23.

60. Holmes MV, Millwood IY, Kartsonaki C, Hill MR, Bennett DA, Boxall R, et al. Lipids, Lipoproteins, and Metabolites and Risk of Myocardial Infarction and Stroke. J Am Coll Cardiol. 2018 Feb 13;71(6):620–32.

61. Paynter NP, Balasubramanian R, Giulianini F, Wang DD, Tinker LF, Gopal S, et al. Metabolic predictors of incident coronary heart disease in women. Circulation. 2018 Feb 20;137(8):841–53.

62. The ARIC Investigators. The Atherosclerosis Risk in Communities (ARIC) Study: design and objectives. . Am J Epidemiol. 1989 Apr;129(4):687–702.

63. Fried LP, Borhani NO, Enright P, Furberg CD, Gardin JM, Kronmal RA, et al. The Cardiovascular Health Study: design and rationale. Ann Epidemiol. 1991 Feb;1(3):263– 76.

64. Feinleib M, Kannel WB, Garrison RJ, McNamara PM, Castelli WP. The Framingham Offspring Study. Design and preliminary data. Prev Med. 1975 Dec;4(4):518–25.

65. Splansky GL, Corey D, Yang Q, Atwood LD, Cupples LA, Benjamin EJ, et al. The Third Generation Cohort of the National Heart, Lung, and Blood Institute’s Framingham Heart Study: design, recruitment, and initial examination. Am J Epidemiol. 2007 Jun 1;165(11):1328–35.

66. Sempos CT, Bild DE, Manolio TA. Overview of the jackson heart study: A study of cardiovascular diseases in african american men and women. Am J Med Sci. 1999 Mar;317(3):142–6.

67. Bild DE, Bluemke DA, Burke GL, Detrano R, Diez Roux AV, Folsom AR, et al. Multi-Ethnic Study of Atherosclerosis: objectives and design. Am J Epidemiol. 2002 Nov 1;156(9):871–81.

68. Benson MD, Eisman AS, Tahir UA, Katz DH, Deng S, Ngo D, et al. Protein-metabolite association studies identify novel proteomic determinants of metabolite levels in human plasma. Cell Metab. 2023 Sep 5;35(9):1646–1660.e3.

69. Ford L, Mitchell M, Wulff J, Evans A, Kennedy A, Elsea S, et al. Clinical metabolomics for inborn errors of metabolism. Adv Clin Chem. 2022;107:79–138.

70. Benedetti E, Chetnik K, Flynn T, Barbieri CE, Scherr DS, Loda M, et al. Plasma metabolomics profiling of 580 patients from an Early Detection Research Network prostate cancer cohort. Sci Data. 2023 Nov 25;10(1):830.

71. Wang N, Ockerman FP, Zhou LY, Grove ML, Alkis T, Barnard J, et al. Genetic Architecture and Analysis Practices of Circulating Metabolites in the NHLBI Trans-Omics for Precision Medicine (TOPMed) Program. BioRxiv. 2025 Feb 3;

72. Friedewald WT, Levy RI, Fredrickson DS. Estimation of the concentration of low-density lipoprotein cholesterol in plasma, without use of the preparative ultracentrifuge. Clin Chem. 1972 Jun;18(6):499–502.

