## Supplementary Text for "Fibrinogen-Associated Plasma Metabolites and Implications for Coagulation, Inflammation, and Vascular Diseases"

**Supplementary Materials**

**Supplementary methods**

*Study descriptions*

The Atherosclerosis Risk in Communities (ARIC) study is a population-based prospective cohort study of cardiovascular disease sponsored by the National Heart, Lung, and Blood Institute (NHLBI). ARIC included 15,792 individuals, predominantly European American and African American, aged 45-64 years at baseline (1987-89), chosen by probability sampling from four US communities. Cohort members completed three additional triennial follow-up examinations, a fifth exam in 2011-2013, a sixth exam in 2016-2017, a seventh exam in 2018-2019, an eighth exam in 2020, and a ninth exam in 2022. The ARIC study has been described in detail previously[1]. Fibrinogen was measured at baseline in the entire ARIC cohort after an 8-hour fasting period. Circulating plasma fibrinogen was measured by the Clauss clotting rate method [2]. Metabolomic profiling was measured at baseline and reported by Metabolon Inc. (Morrisville, NC, USA)

The **Cardiovascular Health Study (CHS)** is a population-based cohort study of risk factors for CHD and stroke in adults ≥65 years conducted across 4 field centers [3,4]. The original predominantly Caucasian cohort of 5,201 persons was recruited in 1989-1990 from random samples of the Medicare eligibility lists; subsequently, an additional predominantly African-American cohort of 687 persons was enrolled for a total sample of 5,888. After an 8-12-h fast, CHS participants underwent phlebotomy by atraumatic venipuncture with a 21-gauge butterfly needle connected to a Vacutainer (Becton Dickinson, Rutherford, NJ) outlet via a Luer adaptor. For fibrinogen determination, an additional citrate-containing tube was processed at 4°C. The study measured fibrinogen levels using the Clauss methods. For metabolomic profiles, measurements were completed at year 7 and reported by the Broad Institute (Boston, MA, USA). Given the small number of African-American participants with metabolite measures at year 7 in CHS (n<70), only measures from participants of European ancestry were used in this analysis.

The **Framingham Heart Study (FHS)** was started in 1948 with 5,209 randomly ascertained participants from Framingham, Massachusetts, US, who had undergone biannual examinations to investigate cardiovascular disease and its risk factors. In 1971, the Offspring cohort (comprising 5,124 children of the original cohort and the children's spouses) and in 2002, the Third Generation (consisting of 4,095 children of the Offspring cohort) were recruited. The methods of recruitment and data collection for the Offspring and Third Generation cohorts have been described [5]. Fibrinogen levels were measured using the Clauss method in the offspring and the third-generation subjects [2], and a modified method of Ratnoff and Menzie in the original cohort subjects [6]. Metabolomic profiling was completed at exam 5 (FHS2) and exam 1 (FHS3) and reported by the Broad Institute (Boston, MA, USA).

The **Multi-Ethnic Study of Atherosclerosis (MESA)** is a cohort study designed to investigate the characteristics of subclinical cardiovascular disease and the risk factors that predict progression to clinically overt cardiovascular disease or progression of the subclinical disease. MESA comprises a diverse, population-based sample of 6,814 asymptomatic men and women aged 45-84. MESA participants are free of clinical cardiovascular disease at baseline. Thirty-eight percent of the recruited participants are Caucasian, 28 percent African-American, 22 percent Hispanic, and 12 percent Asian, predominantly of Chinese descent [7]. Participants were recruited from six field centers across the United States: Wake Forest University, Columbia University, Johns Hopkins University, University of Minnesota, Northwestern University and University of California - Los Angeles.
Fasting blood samples were collected, processed, and stored using standardized procedures. Fibrinogen antigen was measured using the BNII nephelometer (N Antiserum to Human Fibrinogen; Dade Behring Inc., Deerfield, IL). The assay was performed at the Laboratory for Clinical

Biochemistry Research (University of Vermont, Burlington, VT). Intra- and inter-assay analytical coefficients of variation were 2.7% and 2.6%, respectively. Metabolomic profiling was measured at exam 1 and reported by the Broad Institute (Boston, MA, USA).

The **Jackson Heart Study (JHS)** is a cohort study designed to investigate the causes of cardiovascular diseases in African Americans to ultimately improve prophylactic care. JHS builds on the initial Jackson ARIC cohort; a four-center, population-based study including 3,728 African Americans from Jackson, Miss [8]. JHS cohort participants are African American men and women age 35-84 years recruited from the four major counties (Hinds, Madison and Rankin) that make up the Jackson, Miss metropolitan area. Participants are followed up annually with 4 clinical exams in 2000-2004, 2005-2008, 2009-2013 and 2020-2022. Fibrinogen was measured at baseline in the entire JHS cohort after an 8-hour fasting period. Circulating plasma fibrinogen was measured by the Clauss clotting rate method [2]. Metabolomic profiling was measured at visit 1 and reported by the Broad Institute (Boston, MA, USA).

**Supplemental References**

1. Wright, J.D., Folsom, A.R., Coresh, J., Sharrett, A.R., Couper, D., Wagenknecht, L.E., Mosley, T.H., Ballantyne, C.M., Boerwinkle, E.A., Rosamond, W.D., et al. (2021). The ARIC (atherosclerosis risk in communities) study: JACC focus seminar 3/8. J. Am. Coll. Cardiol. *77*, 2939–2959.

2. Clauss, A. (1957). Rapid physiological coagulation method in determination of fibrinogen. Acta Haematol. *17*, 237–246.

3. Cushman, M., Cornell, E.S., Howard, P.R., Bovill, E.G., and Tracy, R.P. (1995). Laboratory methods and quality assurance in the Cardiovascular Health Study. Clin. Chem. *41*, 264–270.

4. Fried, L.P., Borhani, N.O., Enright, P., Furberg, C.D., Gardin, J.M., Kronmal, R.A., Kuller, L.H., Manolio, T.A., Mittelmark, M.B., and Newman, A. (1991). The Cardiovascular Health Study: design and rationale. Ann. Epidemiol. *1*, 263–276.

5. Feinleib, M., Kannel, W.B., Garrison, R.J., McNamara, P.M., and Castelli, W.P. (1975). The Framingham Offspring Study. Design and preliminary data. Prev. Med. *4*, 518–525.

6. Kannel, W.B., Wolf, P.A., Castelli, W.P., and D'Agostino, R.B. (1987). Fibrinogen and risk of

cardiovascular disease. The Framingham Study. JAMA *258*, 1183–1186.

7. Bild, D.E., Bluemke, D.A., Burke, G.L., Detrano, R., Diez Roux, A.V., Folsom, A.R., Greenland, P., Jacob, D.R., Kronmal, R., Liu, K., et al. (2002). Multi-Ethnic Study of Atherosclerosis: objectives and design. Am. J. Epidemiol. *156*, 871–881.

8. Taylor, H.A., Wilson, J.G., Jones, D.W., Sarpong, D.F., Srinivasan, A., Garrison, R.J., Nelson, C., and Wyatt, S.B. (2005). Toward resolution of cardiovascular health disparities in African Americans: design and methods of the Jackson Heart Study. Ethn. Dis. *15*, S6-4.
